# Self-actualization of families with a cerebrovascular disease patient by nurses, and of self-actualization of nurses themselves: An integrative review based on Family Care/Caring Theory

**DOI:** 10.1101/2024.08.09.24311763

**Authors:** Hiroko Ota, Naohiro Hohashi

## Abstract

**Background and purpose:** When caring for patients hospitalized with cerebrovascular disease, a caring phenomenon occurs between the patient’s family and the nurse, and according to Hohashi’s Family Care/Caring Theory, the family and the nurse achieve self-actualization. However, the contents of self-actualization through specific care/caring are unclear. The purpose of this integrative review was to clarify the self-actualization of the nurse as an outcome of family care/caring, and the self-actualization of other individuals (that is, the family) who are supported by nurses.

**Methods:** Conducting a search using Ichushi-Web, CiNii and J-STAGE, which are extensive Japanese literature databases, 1,061 original articles were identified using the keywords “cerebrovascular disease AND nurse.” The quality of the articles was assessed according to the Joanna Briggs Institute critical appraisal checklists. We conducted an integrative review of 11 articles describing transactions between nurses and families according to Toronto and Remington. Self-actualization of the nurse and self-actualization of other individuals were classified as subcategories and then as categories.

**Results:** Of the 11 studies utilized, 10 were qualitative studies and one was a quantitative descriptive study. Eight categories were identified for self-actualization of families with a cerebrovascular patient by nurses, including “Families can realize their hopes through the provision of an environment by nurses.” Three categories of self-actualization of the nurse were identified, including “Nurses can obtain a sense of satisfaction from the family.”

**Conclusion:** During the recovery process after hospitalization for cerebrovascular disease, a caring phenomenon occurs between the patient’s family and nurses, which can be understood using Family Care/Caring Theory. The family’s self-actualization and the nurse’s self-actualization were achieved through reciprocal concern between the two.

## Introduction

Cerebrovascular disease develops suddenly and causes irreversible functional disorders such as consciousness disturbance, paralysis, and speech impairment, leaving serious aftereffects. In Japan, the number of patients hospitalized due to stroke, a typical cerebrovascular disease, reached 123,300 in 2020 [1]. Looking at the number of people transported by ambulance for sudden illnesses [2], circulatory system diseases, including brain diseases and heart diseases, account for 14.7% of the total, and by disease category, emergency transportation is the most common. Patients with cerebrovascular disease who are admitted to the emergency room require immediate treatment, and their symptoms must be monitored in the intensive care unit (ICU) or hospital ward. The families of critically ill patients may suffer emotional, psychological, physical, and social burdens, and in such cases are referred to as postintensive care syndrome families (PICS-F). The prevalence of physiological problems in PICS-F, including depression, anxiety and post-traumatic syndrome, is 20% to 40% at 6 months after ICU admission [3]. Families of patients with cerebrovascular disease experience change and loss, and the family members are easily subject to emotional instability due to the aftereffects of the disease, the risk of recurrence, an uncertain situation, a sense of loss and the burden of taking care of a family member [4].

In other words, care is essential not only for patients with cerebrovascular disease but also for their families/family members. In addition, family care involves family caring [5]. Family Care/Caring Theory (FC2T) [5, 6], a middle-range family nursing theory, was proposed by family nursing researcher/practitioner Naohiro Hohashi in 2013. The FC2T explains nursing and family phenomena in which a relationship is created between the nursing professional and the family, focusing on self-actualization of the nursing professional together with self-actualization of other individuals and the process of establishing a family care/caring relationship.

In FC2T, the noun, family care, and the gerund, family caring, are treated as different concepts. Family care is the acts (practices) of realizing, maintaining, and improving family well-being. On the other hand, family caring is the attitude or mindset of becoming aware of family beliefs and family demands, understanding them, and harnessing them to family care. Family caring involves an alternating relationship of caring for others (family members) and caring for oneself (nurse) simultaneously, through a circular process of transaction between nurses and families (Fig 1). From the nurse’s perspective, supporting the growth, development, and transcendence of the family that is the subject of nursing care, and realizing family well-being (self-actualization of the family, that is, the nurse’s goal). At the same time, it helps nurses achieve their own growth, development, and transcendence, and helps nurses achieve self-actualization. This phenomenon that occurs between nurses and families is the family caring phenomenon [5].

**Fig 1.**
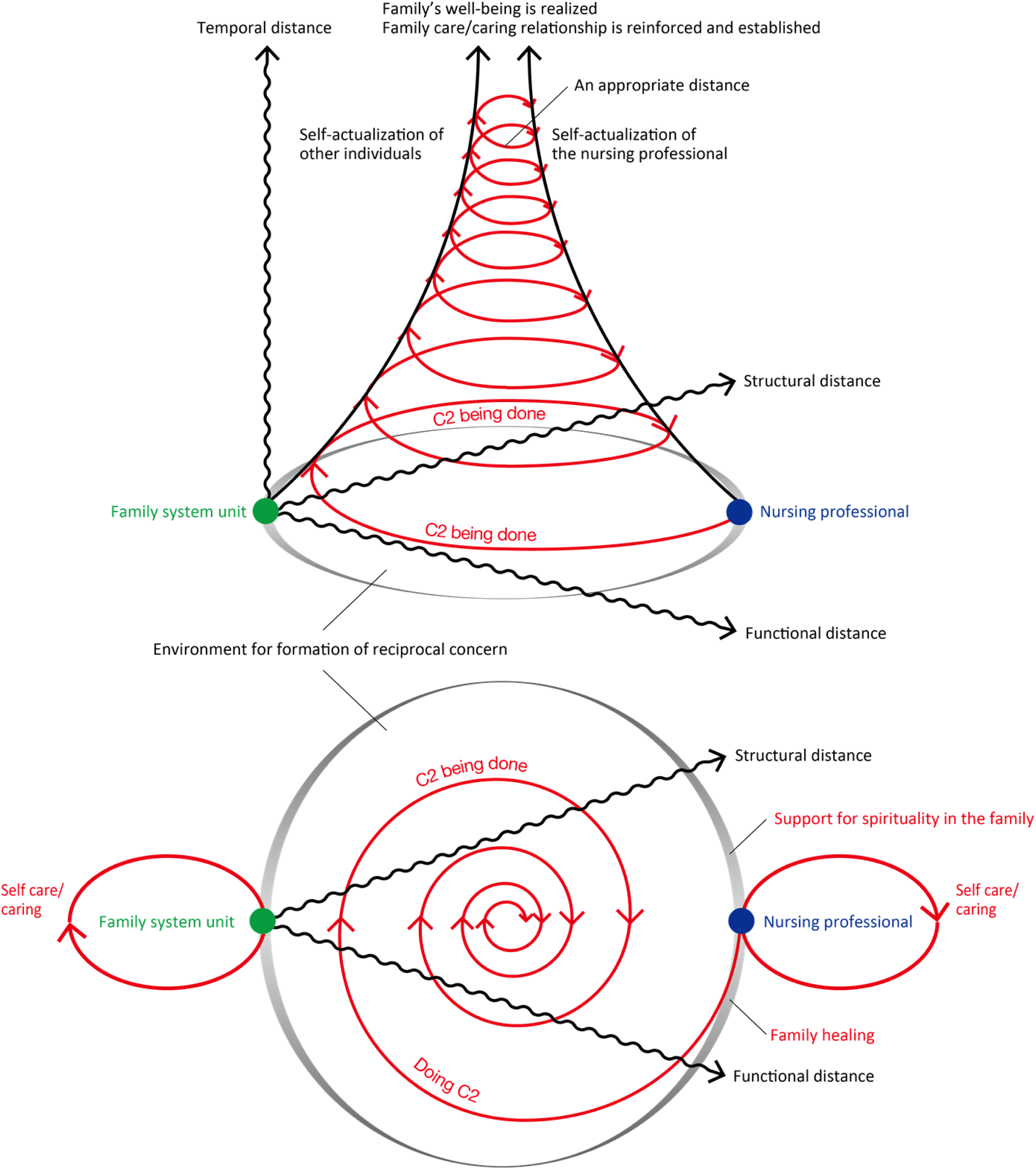
Model diagram of Family Care/Caring Theory (Ver. 2.0). C2 = Care/Caring.

An integrative review of caring [7] revealed that caring behavior leads to increased physical and mental well-being and satisfaction for patients, as well as job satisfaction for nurses. A literature review on the impact of nurses’ caring on families found that families of AIDS patients were able endure their hospitalization, and that home hospice care was highly effective in alleviating family anxiety [8]. When caring for patients with cerebrovascular disease in the ICU, nurses are under considerable stress caused by emotional exhaustion, depersonalization and perceived lack of personal fulfillment [9], making it difficult to practice caring. If smooth communication and transactions between nurses and patients and their families are not realized, it will be difficult for nurses to achieve self-actualization. Family care/caring is also important for alleviating the anxiety of the families of patients with cerebrovascular disease (i.e., for nurses to realize their true selves). Furthermore, it is necessary to materialize self-actualization of other individuals and self-actualization of the nurse.

In this study therefore, we selected literature that describes nursing practices for cerebrovascular disease patients and their families, and focused on the self-actualization of nurses as outcomes of family care/caring, and the self-realization of others supported by nurses (i.e., family self-actualization).

## Methods

### Operational definitions of terms

In this study, we operationally defined terms based on Family Care/Caring Theory (FCCT) [5, 6] as follows:

1. **Family care**: Acts (practices) towards the physical body of the family system unit that are performed with the aim of realizing, maintaining and improving family well-being.
2. **Family caring**: An attitude or mindset for becoming aware of family beliefs and demands, understanding them, and harnessing this understanding to family care.
3. **Family care/caring**: A general term for family care and family caring that maintains unity.
4. **Self-actualization of other individuals**: The process and goal of supporting the growth, development, and transcendence of others in order for them to become who they want to be.
5. **Self-actualization of the nursing professional**: The process and goal of achieving personal growth, development, and transcendence in order to become the person they want to be.

### Search strategy and quality appraisal

An integrative review of this study was conducted based on the instructions of Toronto and Remington [10]. In Japan, scant literature exists on the subject of family caring [11]. We therefore decided to search a wide range of original papers using the keywords “cerebrovascular disease AND nurse” and to extract content related to family care/caring that occurs between families having a patient with cerebrovascular disease and nurses (Fig 2). Three representative literature databases in Japan were utilized: Ichushi-Web, which is medical-related, and CiNii and J-STAGE, which cover a wide range of both literature and science. All articles registered in the databases up to December 2023 were searched, and 1,061 original articles were retrieved, excluding duplicates. Based on the above, 192 articles were selected after applying the following exclusion criteria for title and abstract: no mention of family, no practice by nurses, and no adult patients with cerebrovascular disease. The 192 articles were collected and read closely, and the quantitative and qualitative studies were screened for inclusion and critical appraisal for methodological quality using the Joanna Briggs Institute (JBI) tools. The methodological quality of the studies was assessed by two researchers working independently. In the case of a disagreement between the two reviewers, the item would be discussed and agreement obtained. For the study, in order to assemble a wide range of practical content on family caring, we targeted papers with high or moderate quality ratings.

**Fig 2.**
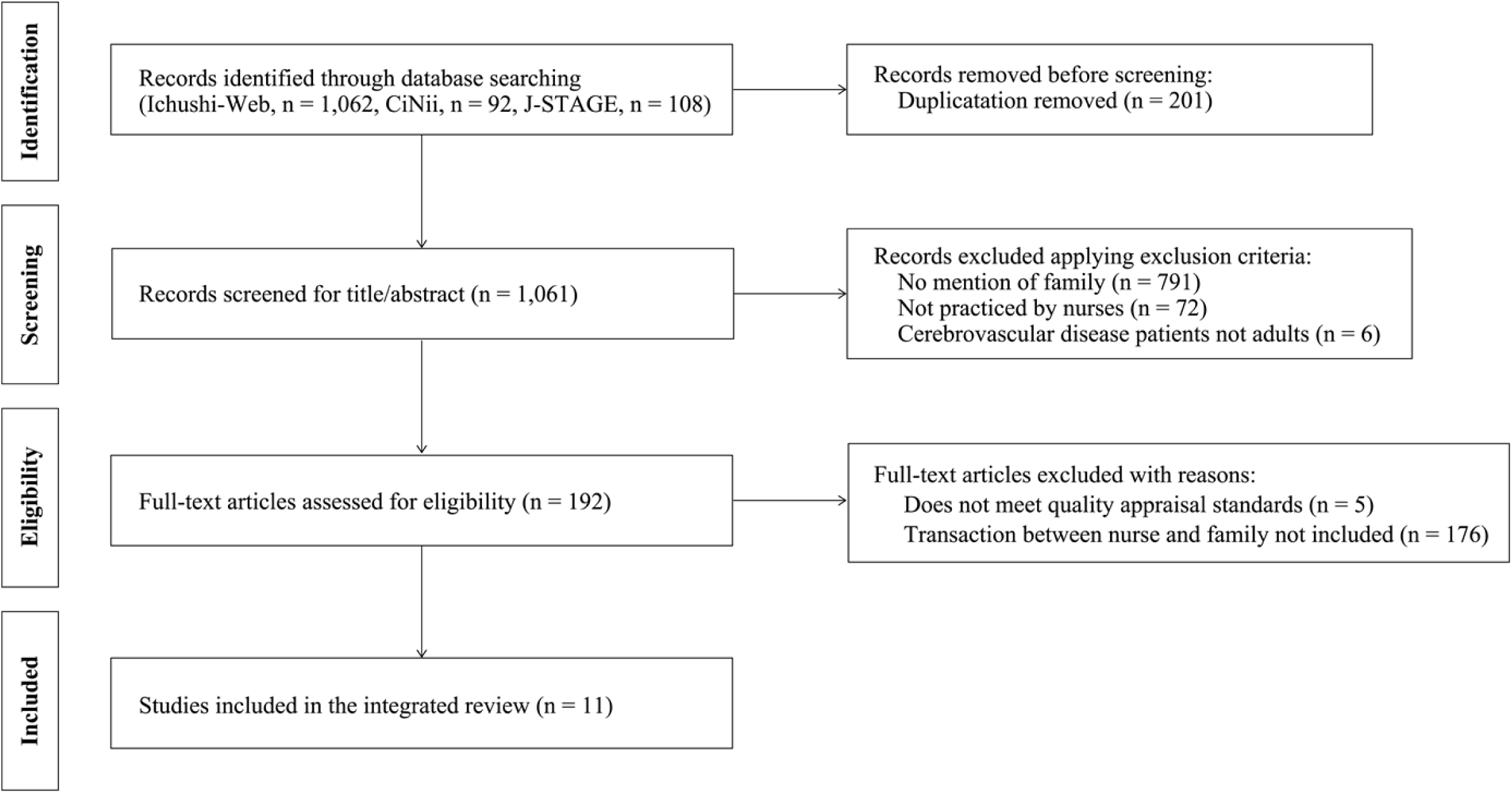
Flow diagram of literature identification and selection.

Specifically, we used the JBI Qualitative Assessment and Review Instrument (JBI-QARI) to appraise the quality of qualitative research [12]. The tool assessed 10 items, including 1) If congruity exists between the stated philosophical perspective and the research methodology; 2) If congruity exists between the research methodology and the research question or objectives; 3) If congruity exists between the research methodology and the methods used to collect data; 4) If congruity exists between the research methodology and the representation and analysis of data; 5) If congruence exists between the research methodology and the interpretation of results; 6) If the researcher is located culturally or theoretically; 7) If influence of the researcher on the research, and vice-versa, is adequately addressed; 8) If participants and their voices are represented; 9) If ethical approval by an appropriate body was obtained; and 10) If a relationship of conclusions to analysis, or interpretation of the data, was established. Each item was assessed as “Yes,” “No,” “Unclear” or “Not applicable,” with the total number of “Yes” items evaluated as follows: low (0-4 items), moderate (5-7 items) and high (8-10 items).

Additionally, we used the JBI Checklist for Analytical Cross-Sectional Studies to appraise the quality of quantitative research [13]. This tool assessed 8 items, including 1) If the criteria for inclusion in the sample were clearly defined; 2) If the study subjects and the setting were described in detail; 3) If the exposure was measured in a valid and reliable way; 4) If objective, standard criteria were used for measurement of the condition; 5) If confounding factors were identified; 6) If strategies to deal with confounding factors were stated; 7) If the outcomes were measured in a valid and reliable way; and 8) If appropriate statistical analysis was used. Each item was assessed as “Yes,” “No,” “Unclear” or “Not applicable,” with the total number of “Yes” items evaluated as follows: low (0-4 items), moderate (5 or 6 items) and high (7 or 8 items). Five papers that did not meet the JBI criteria of high or moderate were excluded. Furthermore, based on the Family Care/Caring Theory, 176 papers that did not include interactions between nurses and families were excluded, leaving 11 papers for analysis [14–24].

### Data analysis procedure and trustworthiness

After carefully perusing the 11 papers, we identified self-actualization of the nursing professionals and self-actualization of other individuals from descriptions of family care/caring expressed in the results, and wrote them in concise sentences without sacrificing meaning. This was then encoded. In addition to the codes, the literature was organized using Garrard’s matrix method [25] based on the article title, research purpose, stage classification of patients with cerebrovascular disease, and JBI Qualitative Assessment and Review Instrument or JBI Checklist for Analytical Cross-Sectional Studies scores. The codes were subsequently classified and subcategories were named, taking into consideration the commonality and specificity of the content. After that, we found commonalities and differences from the subcategories and named them as categories by increasing the level of abstraction [26]. The categories were deductively classified into two main categories: self-actualization of other individuals and self-actualization of the nursing professional [27].

All analyses were reviewed by two family nursing researchers until a consensus was reached. At the same time, we received repeated opinions from seven researchers to ensure the trustworthiness of the analysis.

### Ethical consideration

This research did not require approval from an institutional review board as neither humans nor animals were involved. In conducting the research, we complied with research ethics and clearly indicated sources to avoid problems of copyright infringement. The written content of the paper was then closely read and written accurately so that the authors’ intent and meaning were not compromised.

## Results

### Details and characteristics of the included studies

Details and characteristics of the included studies are indicated in Table 1. The purpose of the research was to clarify nurses’ practices, their processes, and their relationships with patients and their families, and others. Of the 11 papers, 10 were qualitative studies and one was a quantitative descriptive study using the Delphi method. As for stage the classification of patients with cerebrovascular disease in the 11 papers, six were acute stage and five were convalescent stage.

**Table 1.**
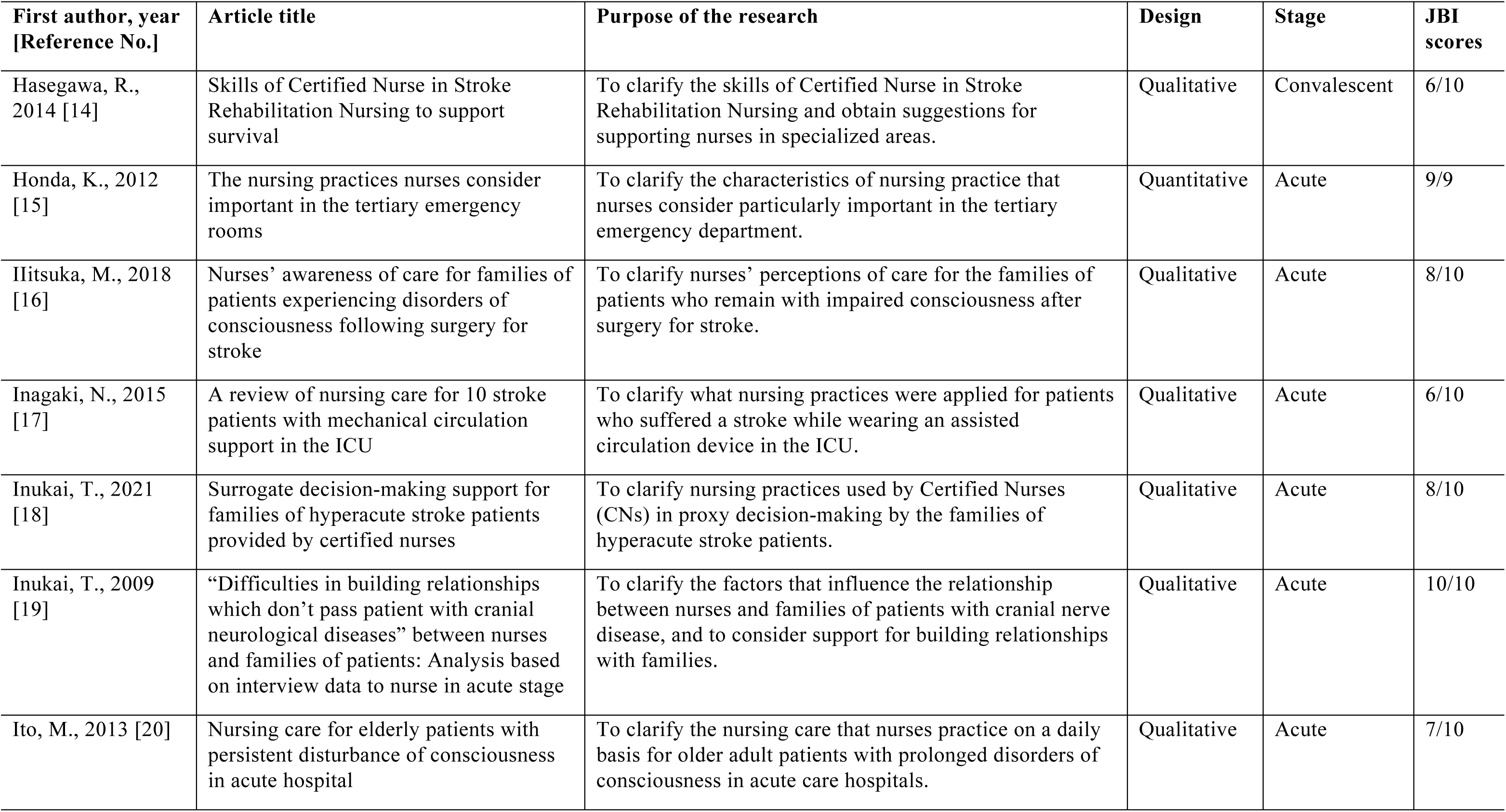
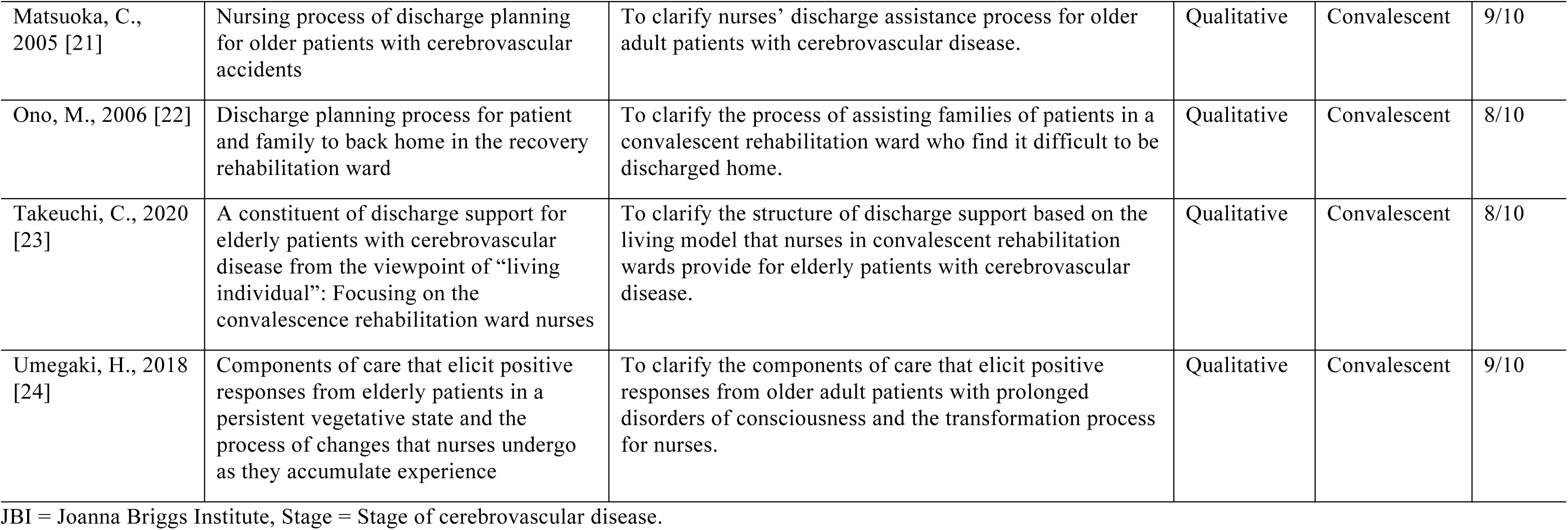
Details and characteristics of the included studies.

| **First author, year**<br>**[Reference No.]** | **Article title** | **Purpose of the research** | **Design** | **Stage** | **JBI**<br>**scores** |
| --- | --- | --- | --- | --- | --- |
| Hasegawa, R.,<br>2014 [14] | Skills of Certified Nurse in Stroke Rehabilitation Nursing to support survival | To clarify the skills of Certified Nurse in Stroke Rehabilitation Nursing and obtain suggestions for supporting nurses in specialized areas. | Qualitative | Convalescent | 6/10 |
| Honda, K., 2012<br>[15] | The nursing practices nurses consider important in the tertiary emergency rooms | To clarify the characteristics of nursing practice that nurses consider particularly important in the tertiary emergency department. | Quantitative | Acute | 9/9 |
| Iiitsuka, M., 2018<br>[16] | Nurses' awareness of care for families of patients experiencing disorders of consciousness following surgery for stroke | To clarify nurses' perceptions of care for the families of patients who remain with impaired consciousness after surgery for stroke. | Qualitative | Acute | 8/10 |
| Inagaki, N., 2015<br>[17] | A review of nursing care for 10 stroke patients with mechanical circulation support in the ICU | To clarify what nursing practices were applied for patients who suffered a stroke while wearing an assisted circulation device in the ICU. | Qualitative | Acute | 6/10 |
| Inukai, T., 2021<br>[18] | Surrogate decision-making support for families of hyperacute stroke patients provided by certified nurses | To clarify nursing practices used by Certified Nurses (CNs) in proxy decision-making by the families of hyperacute stroke patients. | Qualitative | Acute | 8/10 |
| Inukai, T., 2009<br>[19] | "Difficulties in building relationships which don't pass patient with cranial neurological diseases" between nurses and families of patients: Analysis based on interview data to nurse in acute stage | To clarify the factors that influence the relationship between nurses and families of patients with cranial nerve disease, and to consider support for building relationships with families. | Qualitative | Acute | 10/10 |
| Ito, M., 2013 [20] | Nursing care for elderly patients with persistent disturbance of consciousness in acute hospital | To clarify the nursing care that nurses practice on a daily basis for older adult patients with prolonged disorders of consciousness in acute care hospitals. | Qualitative | Acute | 7/10 |
| Matsuoka, C., 2005 [21] | Nursing process of discharge planning for older patients with cerebrovascular accidents | To clarify nurses' discharge assistance process for older adult patients with cerebrovascular disease. | Qualitative | Convalescent | 9/10 |
| Ono, M., 2006 [22] | Discharge planning process for patient and family to back home in the recovery rehabilitation ward | To clarify the process of assisting families of patients in a convalescent rehabilitation ward who find it difficult to be discharged home. | Qualitative | Convalescent | 8/10 |
| Takeuchi, C., 2020 [23] | A constituent of discharge support for elderly patients with cerebrovascular disease from the viewpoint of "living individual": Focusing on the convalescence rehabilitation ward nurses | To clarify the structure of discharge support based on the living model that nurses in convalescent rehabilitation wards provide for elderly patients with cerebrovascular disease. | Qualitative | Convalescent | 8/10 |
| Umegaki, H., 2018 [24] | Components of care that elicit positive responses from elderly patients in a persistent vegetative state and the process of changes that nurses undergo as they accumulate experience | To clarify the components of care that elicit positive responses from older adult patients with prolonged disorders of consciousness and the transformation process for nurses. | Qualitative | Convalescent | 9/10 |

JBI = Joanna Briggs Institute, Stage = Stage of cerebrovascular disease.

### Self-actualization of other individuals

Eight subcategories and 18 categories were extracted for self-actualization of other individuals (Table 2). During the course of a family member developing and recovering from cerebrovascular disease, family caring provided by nurse as an expert could help achieve the family self-actualization. Below, categories are indicated inside quotation marks.

**Table 2.**
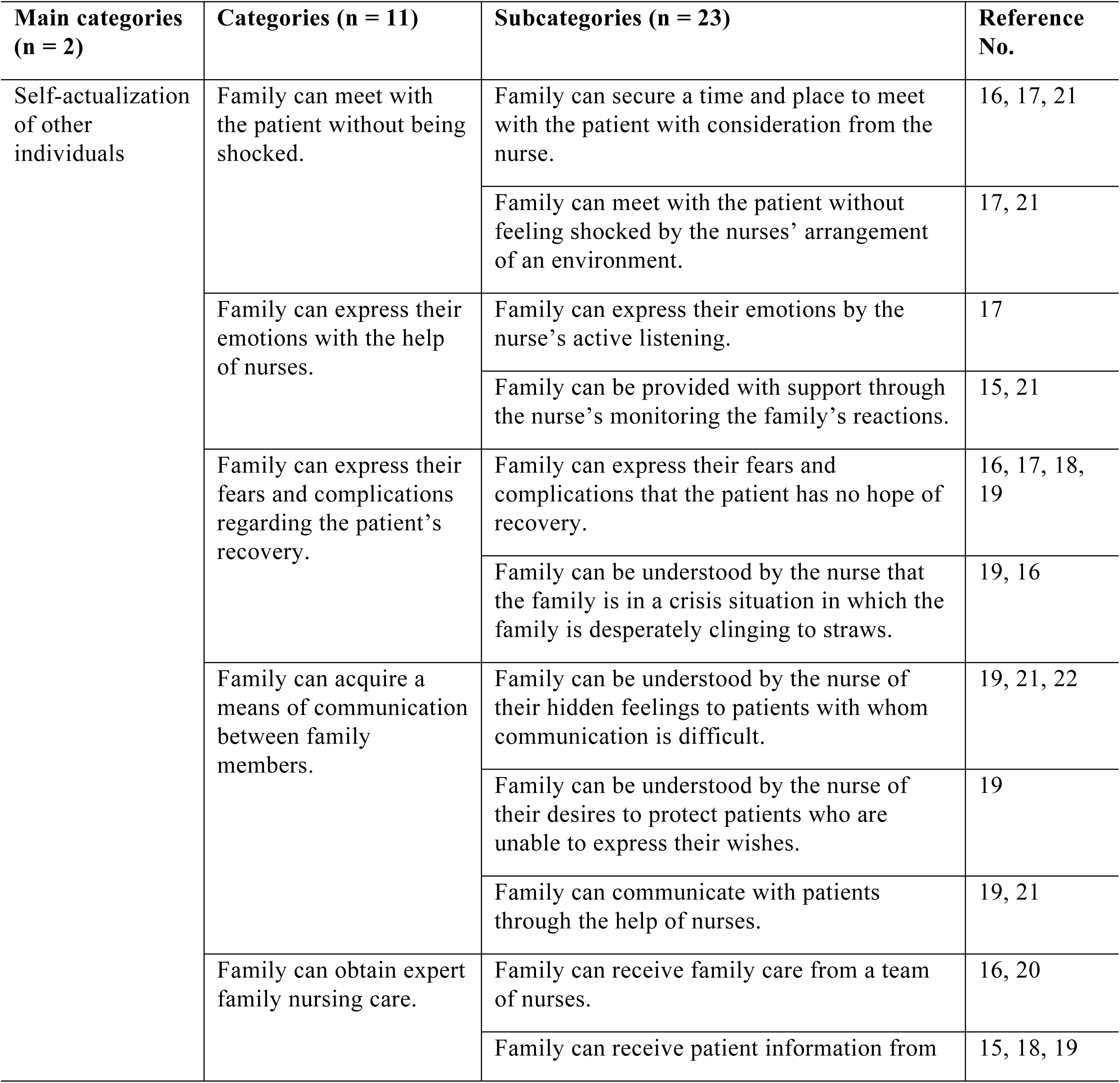
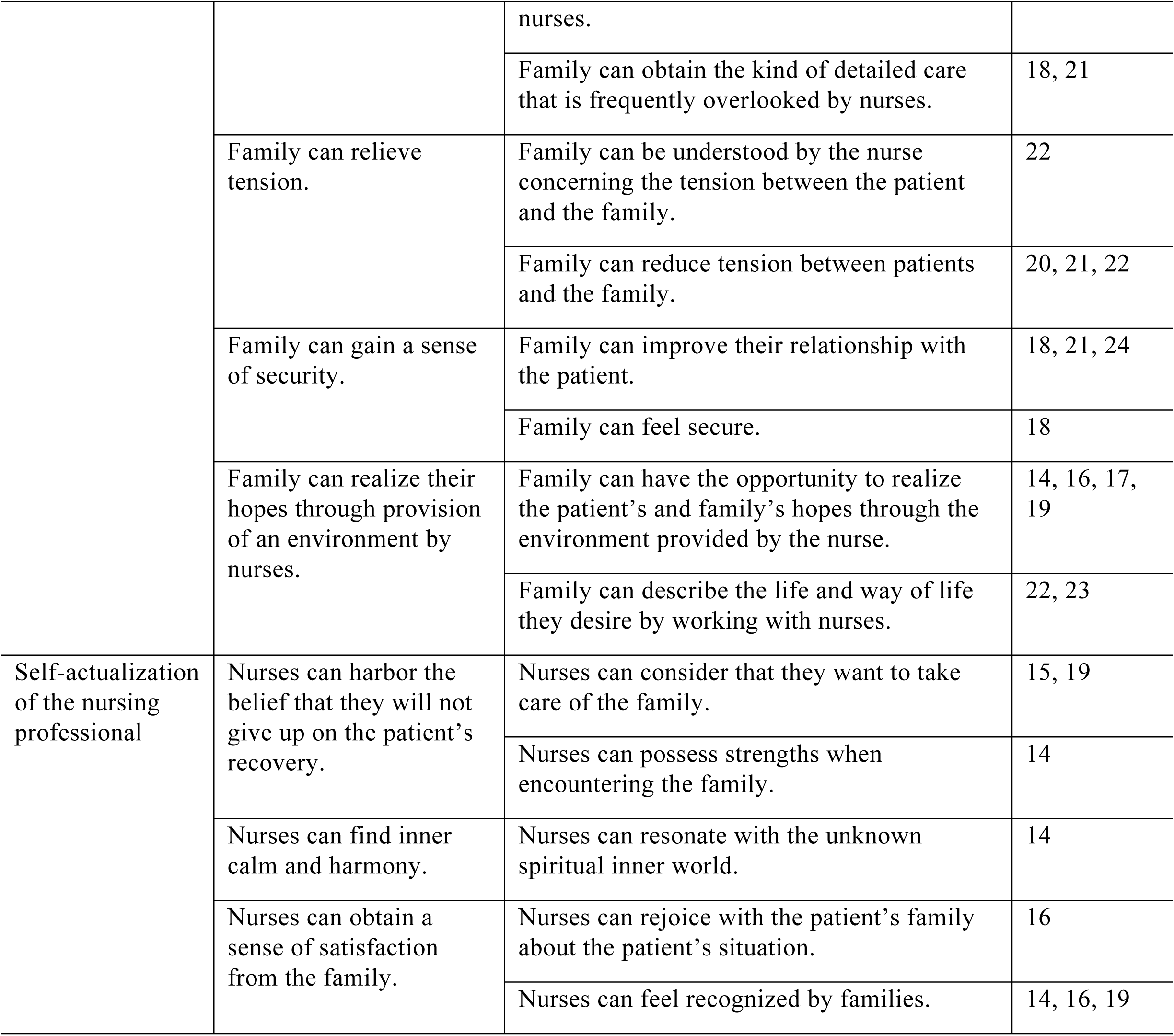
Categories of self-actualization of families with a cerebrovascular disease patient by the nurse, and of self-actualization of the nurse.

| Main categories<br>(n = 2) | Categories (n = 11) | Subcategories (n = 23) | Reference<br>No. |
| --- | --- | --- | --- |
| Self-actualization<br>of other<br>individuals | Family can meet with<br>the patient without being<br>shocked. | Family can secure a time and place to meet<br>with the patient with consideration from the<br>nurse. | 16, 17, 21 |
|  |  | Family can meet with the patient without<br>feeling shocked by the nurses' arrangement<br>of an environment. | 17, 21 |
|  | Family can express their<br>emotions with the help<br>of nurses. | Family can express their emotions by the<br>nurse's active listening. | 17 |
|  |  | Family can be provided with support through<br>the nurse's monitoring the family's reactions. | 15, 21 |
|  | Family can express their<br>fears and complications<br>regarding the patient's<br>recovery. | Family can express their fears and<br>complications that the patient has no hope of<br>recovery. | 16, 17, 18,<br>19 |
|  |  | Family can be understood by the nurse that<br>the family is in a crisis situation in which the<br>family is desperately clinging to straws. | 19, 16 |
|  | Family can acquire a<br>means of communication<br>between family<br>members. | Family can be understood by the nurse of<br>their hidden feelings to patients with whom<br>communication is difficult. | 19, 21, 22 |
|  |  | Family can be understood by the nurse of<br>their desires to protect patients who are<br>unable to express their wishes. | 19 |
|  |  | Family can communicate with patients<br>through the help of nurses. | 19, 21 |
|  | Family can obtain expert<br>family nursing care. | Family can receive family care from a team<br>of nurses. | 16, 20 |
|  |  | Family can receive patient information from nurses. | 15, 18, 19 |
|  |  | Family can obtain the kind of detailed care that is frequently overlooked by nurses. | 18, 21 |
|  | Family can relieve tension. | Family can be understood by the nurse concerning the tension between the patient and the family. | 22 |
|  |  | Family can reduce tension between patients and the family. | 20, 21, 22 |
|  | Family can gain a sense of security. | Family can improve their relationship with the patient. | 18, 21, 24 |
|  |  | Family can feel secure. | 18 |
|  | Family can realize their hopes through provision of an environment by nurses. | Family can have the opportunity to realize the patient's and family's hopes through the environment provided by the nurse. | 14, 16, 17, 19 |
|  |  | Family can describe the life and way of life they desire by working with nurses. | 22, 23 |
| Self-actualization of the nursing professional | Nurses can harbor the belief that they will not give up on the patient's recovery. | Nurses can consider that they want to take care of the family. | 15, 19 |
|  |  | Nurses can possess strengths when encountering the family. | 14 |
|  | Nurses can find inner calm and harmony. | Nurses can resonate with the unknown spiritual inner world. | 14 |
|  | Nurses can obtain a sense of satisfaction from the family. | Nurses can rejoice with the patient's family about the patient's situation. | 16 |
|  |  | Nurses can feel recognized by families. | 14, 16, 19 |

Nurses gave consideration to listen to family other than at bedside, and even when a cerebrovascular patient had been admitted emergently and visits were restricted, they gave consideration to secure the visiting time and place that the family desired. Nurses created an environment in which the family could participate in patient care such as grooming and others during visits, and by keeping the patient’s hands clean so that visiting family members could hold the patient’s hands. As a result, “the family could meet with the patient without being shocked.”

The nurse made an effort to understand the family’s reaction and the family’s feelings of grief over the lack of treatment options for the patient, and to perform active listening to the family. Through this relationship, “the family was able to express their emotions with the help of nurses.”

Faced with the condition of the family member who had developed cerebrovascular disease, the family felt fear, conflict, and regret that the patient’s recovery was hopeless. In addition, the family’s desire by the nurses to know the patient’s medical condition and prognosis could be understood. Through these transactions with nurses, “families could express their fears and complications regarding the patient’s health.” The family’s concealed feelings for the patient was understood by the nurse, even though it was difficult to communicate with the patient due to decreased consciousness and language impairment due to cerebrovascular disease. Additionally, families were able to get nurses to understand their desire to protect patients who were unable to express their wishes, and by receiving bridging support from nurses, “families were able to acquire a means of communication between family members.”

The family was able to receive professional family care from a team of nurses. This means that even when the patient’s treatment and progress were specialized, the family could easily understand the patient’s treatment and progress, and by being able to receive comprehensive family care that might easily be overlooked, “the family could obtain expert family nursing care.”

As the patient moved out of the acute phase and began to recover, the family and patient were conflicted about the place where the patient would be discharged from the hospital and what direction to take in the future. The family was understood by nurses that family members interacted with each other while concealing their feelings, and was provided opportunities for discussion between the patient and family. As a result, “family could relieve tension.”

The nurse intervened between family members by providing caring, and the family felt that their relationship with the patient had improved, and that they had developed a sense that “the family could gain a sense of security.” Even when the family was unable to communicate with the patient, the family felt that nurses shared family hopes and the hopes the patient had harbored before symptoms developed, and incorporated them into the patient’s care.

Furthermore, by exploring the patient’s meaning in life with family members, the family worked with nurses to describe the image of the life that both the patient and family would like, which led to “family could realize their hopes through the provision of an environment by nurses.”

### Self-actualization of the nursing professional

Five subcategories and three categories were extracted for self-actualization of the nursing professional (Table 2). Below, categories are indicated inside double quotation marks. By being happy with the family of the patient with cerebrovascular disease and feeling recognized by the family, the nurse was able “to obtain a sense of satisfaction from the family.” In addition, through the experience of resonance with an unknown spiritual world through the practice of family caring, nurses were able “to find inner calm and harmony,” and felt a sense of inner self-growth. Underlying this were nurses’ beliefs that the words of family members were important, their desire to take care of the family, and possession of strengths when encountering the family, and that “nurses harbor the belief that they will not give up on the patient’s recovery.” The nurses had such a belief and practiced family caregiving at the same time as caring for patients with cerebrovascular disease who had difficulty communicating. While attending to the feelings of the family members, the nurses experienced a spiritual inner world and felt as if their own energy was amplified.

## Discussion

### Self-actualization of other individuals

Cerebrovascular disease often develops suddenly, putting families in a critical state not unlike the patient. The family can meet with the patient without feeling shocked because they feel the patient is receiving the best possible care, such as by seeing the patient well-groomed when visiting and participating in the patient’s care, which can be thought of as self-actualization of the family. When a family feels that a patient in a life-threatening situation is receiving the best possible treatment and care, their suffering is alleviated [5]. Families with cerebrovascular patients achieved self-actualization through nurses’ family caring from their emergency hospitalization to the recovery process. Families are able to express their emotions with the help of nurses and express their fears and complications regarding the patient’s recovery. Family members who see the sudden onset of symptoms become upset, realize that the patient’s life is in danger, may suffer from sequelae, and feel helpless and regretful that the patient has changed from before the onset of symptoms [28]. Emotional involvement may also affect family functioning and lead to psychological trauma [29]. We believe that nurses’ assistance in being able to express the family’s emotions provides an opportunity for the family to receive psychological support [30].

Cerebrovascular disease is a pathological condition caused by damage to the cranial nerves due to cerebral infarction or cerebral hemorrhage, resulting in aftereffects such as impaired consciousness, language impairment, and motor paralysis, leading to a decline in family functioning due to decreased interaction with family members as a result of impaired communication [29]. Even in a situation where family interaction with the patient is reduced, nurses are able to act as a mediator, allowing families to acquire a means of communication between family members and obtain expert family nursing care. The family is in an uncertain situation, unable to understand the patient’s treatment and future course. Considering the family situation, it is important for nurses to support the patient and family by using verbal, nonverbal, and descriptive means so that the family can communicate with the patient. Because obstructed communication can affect the relationship between patients and families [31], nurses provide information to families by modifying specialized contents into language that families can easily understand [32]. In addition, through nurses’ understanding that the family wishes to receive truthful information, the family is able to gain an opportunity to communicate with the patient. When families are confused and desperate, a sensitive nurse who spends time with them and listens to them can be the best help [30]. We believe that nurses, as professionals who can provide delicate care to families, are important for the family’s self-actualization.

As the patient recovers, the family is relieved that the patient is out of the acute phase, but at the same time, are still uncertain about the aftereffects and what kind of life they will need to lead in the future, and they are trying to determine how to live their lives henceforth [33]. Uncertainty about the disease state is highest immediately after the onset of the disease, and although it decreases, it rises again, and the family continues to feel uneasy about the unknown future [34]. Because of this uncertainty, the family becomes anxious about decision-making and are hesitant when dealing with the patient’s feelings. Tensions between family members can be alleviated by being regulated by nurses. Furthermore, the family feels that relationships between family members have improved, which leads to the family’s self-actualization through the gaining of a sense of security. Even in situations where family members are unable to communicate with the patient, nurses provide care that incorporates the patient’s wishes and preferences, and nurses adjust the environment to portray the patient’s purpose in life and the image of the life desired by the family, enabling self-actualization, such as through realization of their hopes, to be achieved. In order to build a relationship of trust for the future of the family, nurses practice family caring, starting from an interest in the patient’s family and exploring the wishes of the family [30]. Attentive reassurance indicates availability and a more hopeful outlook [35]. The nurse’s attitude of searching for the family’s desired way of life leads to self-actualization of the family that attends to the unconscious patient, through such ways as gaining detailed reassurance and achieving the family’s wishes. In addition, nurses’ support produces a transaction by which patients and nurses work together to create meaning, and when they find a path to recovery, they promote self-actualization in patients [36]. The constructive philosophy of family caring proposed by Hohashi incorporates the key factors that make up family caring, that is, concern, knowledge, trust, and hope [5]. The results suggest that not only patients but also families experience the philosophy of family caring through family caring practices by nurses, leading to family self-actualization.

### Self-actualization of the nursing professional

Because cerebrovascular disease is characterized by changes in condition and uncertainty [4], not only patients and families, but also nurses may feel anxious and wonder whether they should provide care actively. In this way the nurse’s feeling of being accepted by the family may lead to a sense of satisfaction from the family as well as finding inner calm and harmony. At the root of this is nurses’ belief that the words of families are important, their desire to place value on families, and their possession of strengths when they meet families, as a result nurses harbored the belief that they will not give up on the patient’s recovery.

Self-actualization is extremely important for nurses as professionals, and they have an internal drive to excel at their work [37]. It can be seen that the nurses have a strong desire for the patient’s recovery, recognize that the presence of the family is important for that purpose, and practice family caring. Also, during interactions with families, nurses experienced a spiritual inner world and felt that their own energy was amplified. According to Watson, when the nurse enters into the other’s experience new phenomena, referred to as Caritas Field, is created that is greater than that of two persons; thus another person enters into the nurse’s experience. And the union of feelings can potentiate self-healing, discovery of power and control and contribute to another person finding meaning in their existence [38]. Hohashi explains that spiritual dimension is the realm where something fundamental exists, which gives a person meaning and purpose in life, and can be understood from relationships and time [5]. While the patient is in an uncertain situation, nurses and their families deepen their relationship with each other over the course of the time they care for the patient, promoting mutual understanding, recognizing that they are necessary, and fostering self-growth, that is, inner self-growth. It is possible that they feel that their personal growth is expanding.

### Storyline based on Family Care/Caring Theory

In Hohashi’s Family Care/Caring Theory (FC2T) (Figure 1), reinforcing and establishing family care/caring relationships is understood in terms of three distances: structural distance, functional distance, and temporal distance [5, 6]. In other words, nurses provide family care/caring to families, and in the process of mutual care/caring, the structural distance and functional distance between nurses and families become closer in the environment for formation of reciprocal concern, and the family’s well-being will be realized. Based on the categories clarified in this study, the storyline of the relationship between nurses and families based on temporal distance is as follows: The starting point for care/caring by nurses begins when a cerebrovascular patient is admitted to the hospital. Owing to the care/caring of the nurses, the family is able to meet with the patient without being shocked and is able to express their emotions with the help of the nurse. Through the process of nurses interacting with families as professionals, families were able to express their fears and complications regarding the patient’s recovery, and the nurse’s role as a bridge enabled families to acquire a means of communication between family members. As the patient’s condition improved, the family was able to relieve tension, gain a sense of security, and realize their hopes through provision of an environment by the nurses. For nurses on the other hand, over the passing of time they achieved self-actualization such as observing a sense of satisfaction from the family and finding inner calm and harmony. At the basis of this was the belief that nurses will not give up on the cerebrovascular patient’s recovery.

### Limitations

Because this study analyzed Japanese literature, the contents regarding self-actualization of the nursing professional and self-actualization of other individuals may differ depending on culture. In addition, there are scant examples of literature that study transactions between entire families and nurses, or self-actualization of families with a cerebrovascular disease patient by nurses, and of self-actualization of nurses themselves. Consequently it is difficult to draw general conclusions from the results of this study. In the future, we intend to study the building of relationships between nurses and families and the self-actualization of both parties based on practical situations of family care/caring that nurses practice with families, and will need to clarify specifically what kinds of transactions occur.

## Conclusion

After a patient is hospitalized due to cerebrovascular disease, the patient’s family receives care/caring from nurses during the recovery process, gaining self-actualization such as by the obtaining of a sense of security. Nurses, on the other hand, obtained care/caring from the patients’ families and gained self-actualization such as through observing the family’s satisfaction. This phenomenon of care/caring can be understood using Family Care/Caring Theory, and a relationship can be established through reciprocal concerns of care/caring between nurses and families.

## Data Availability

All relevant data are within the manuscript and its Supporting Information files.

## Acknowledgments

Our laboratory members’ discussions in the data analysis were instrumental in ensuring its trustworthiness. For their conscientious efforts, the authors would like to express our deepest thanks and appreciation.

